# Telehealth Care for Adolescents and Young Adults with HIV: Practices and Perspectives from a National Mixed-Methods Study of U.S. HIV Care Providers

**DOI:** 10.1101/2025.08.11.25333436

**Authors:** E. A. Barr, R.P. Momin, Q. Qian, R. Beach, M. Lingwall, M. E. Paul, H. Armitage, H. Wu, T. P. Giordano

**Author notes:** Corresponding author: Emily Anne Barr, PhD, RN, CPNP-PC, CNM, ACRN, FACNM, FAAN University of Buffalo, School of Nursing 101 Wende Hall, 3435 Main St, Buffalo, NY 14214.

## Abstract

**Background:** Youth living with HIV (YWH) face persistent barriers to care engagement and viral suppression. Telehealth expanded during COVID-19, offering new possibilities for access, but provider perspectives on its use with YWH are underexplored.

**Methods:** A national mixed-methods survey captured experiences of 156 HIV care providers. Quantitative data were analyzed using descriptive and inferential statistics; qualitative data were analyzed using reflexive thematic analysis.

**Results:** Most providers (88.5%) believed telehealth would continue to play a role in HIV care. Reported benefits included improved access (80.7%) and workflow efficiency (64.7%). However, providers were significantly less likely to discuss sensitive topics “often” via telehealth compared to in-person care, including mental health (81.4% vs. 92.9%, p = 0.0008) and sexual health (77.9% vs. 93.6%, p < 0.0001).

**Conclusions:** While telehealth improves access for YWH, hybrid care models and targeted provider training are needed to address privacy, communication, and trust when discussing sensitive health concerns.

## Introduction

The HIV epidemic continues to be a significant and persistent global public health crisis that disproportionately impacts adolescents and young adults. ^1^ Worldwide, approximately 5 million youth aged 15-24 have HIV, ^2^ and this population faces heightened risks of poor health outcomes and continued transmission. ^3^ Despite advances in HIV prevention and treatment, various risk factors, including limited access to healthcare, stigma, and lack of awareness, underscore the critical need for targeted interventions and innovative care strategies adapted to the special needs of youth with HIV (YWH). ^4^

The onset of the COVID-19 pandemic caused significant disruptions to healthcare delivery systems globally, prompting a rapid shift to telehealth services, including in the care of HIV. ^5^ While this transition presented challenges, it also created a pivotal opportunity to reimagine how HIV care is delivered to youth. Telehealth, including synchronous audio and video consultations and audio-only interactions, quickly emerged as an essential tool for maintaining continuity of care when traditional in-person services were unavailable or restricted.^6^

The transition to telehealth was supported by federal policy changes and clinical guidelines, enabling telehealth’s seamless integration into HIV care. ^7^ Telehealth has emerged as a preferred modality among YWH, aligning with their communication styles and lifestyles. Its convenience, privacy, and accessibility offer promising pathways to improve engagement and retention in care, improving access, supporting adherence, and, in some cases, contributing to better viral suppression outcomes. ^6^ In the United States, 99% of surveyed HIV providers offered telehealth services, with 47% of visits delivered through this modality. Among patients, 57% expressed a preference for telehealth over in-person visits. ^8^ As telehealth becomes increasingly embedded in care models, understanding its specific impact on populations like YWH and their providers remains essential.

Despite the benefits and quick adoption of telehealth, challenges emerged, including limited access to technology, private space, and timely laboratory testing, barriers that were particularly pronounced among YWH. ^8^ In metropolitan DC, a study focusing on predominantly Black children and YWH, lower rates of laboratory testing with telehealth visits underscored the need for improved infrastructure to support comprehensive remote care. ^8^ The reduction of in-person interactions from the uptake in telehealth has raised concerns about the potential impact on care quality and the ability to perform necessary physical assessments. ^9^ Boshara (2022) highlighted the digital divide as a critical barrier to telehealth access, particularly among marginalized YWH, suggesting the need for targeted interventions to address these disparities. ^10^

Healthcare providers play a pivotal role in the successful integration of telehealth into HIV care models. While the benefits of telehealth for patients, particularly YWH, are well-documented, the perceptions, challenges, and experiences of the providers who deliver this care are equally critical to its long-term success. Understanding these perspectives is essential to optimizing telehealth to meet the unique needs of this vulnerable population. Enhancing provider training and telehealth infrastructure may further strengthen care delivery, but additional research is needed to fully understand and support these dynamics. ^6,11^

While telehealth is increasingly used in HIV care, most existing research emphasizes patient-level outcomes, with limited understanding of the perspectives of providers delivering this care, particularly to YWH. ^4,6^ This paper examines HIV healthcare provider perspectives on telehealth use when caring for YWH. By exploring provider attitudes, satisfaction levels, and perceptions of telehealth’s benefits and barriers, we aim to uncover critical insights that can guide the effective integration of telehealth into HIV care models. Understanding the factors that influence provider buy-in and utilization is essential for optimizing care delivery and ensuring sustained patient engagement.

## Methods

This study employs a mixed methods embedded design, in which qualitative data play a supporting role within a primarily quantitative study. ^12^ This method was utilized to contextualize and provide deeper insights into quantitative findings. ^13^ This mixed-methods study aimed to quantitatively describe telehealth experiences and to better understand the benefits and challenges of telehealth use in the care of YWH, through soliciting the qualitative perspectives of a diverse group of healthcare providers.

The survey instrument used in this study was developed by adapting existing surveys (Table 1) and questions derived from existing literature on telehealth care in YWH. ^14–28^ To gain deeper insight into the experiences and perceptions of healthcare providers, the survey asked providers to identify the benefits and barriers to telehealth use, best practices, strategies to build and maintain patient-provider trust, and ways to use telehealth to enhance HIV care for YWH. The survey also collected demographics, provider type, and specific practices and challenges in providing telehealth services to this population. Qualitative data were collected concurrently with the quantitative phase, serving to enhance understanding of key patterns emerging from numerical findings. ^12^ Open-ended or free-text questions were incorporated to allow respondents to share experiences and perspectives beyond the scope of the predefined response options.

**Table 1.** Telehealth Surveys Used in Survey Design.

| Title of Tool/Instrument | Citation | Survey information |
| --- | --- | --- |
| Baylor College of Medicine Telehealth Survey | Guajardo et al., 2022 | A cross-sectional study evaluating the telehealth experiences of providers (n=157) caring for Latinx people with HIV, includes survey instrument developed from validated items. |
| Swedish Primary Care Physician's Survey | Pikkemaat et al., 2021 | Qualitative study exploring provider (n=198) intentions to use telehealth before and after the COVID-19 pandemic; includes novel survey items tested for construct validity, internal validity, and reliability. |
| Massachusetts General Hospital Telehealth provider Survey under "Technical Issues" | Donelan et al., 2019 | Qualitative study of provider (n=74) and patient (n=426) perceptions of telehealth experiences compared to in-person medical care, includes over 25 validated Likert measures. |

**Note:** Includes instruments that were adapted for use in the survey for this study.

The study included US HIV care providers, specifically Registered Nurses (RNs), Advanced Practice Providers (APPs; ie. Advanced Practice Nurses (APRNs) and Physician Assistants (PAs)) and Physicians (MD/DOs), who provide care for YWH ages 16-29 years of age. Inclusion criteria required providers be 18 years of age or older, able to read and write in English, and must have provided care in a clinical setting for YWH for 2 or more years. Participants were not required to have previous telehealth experience to respond to the survey. This criterion ensures a comprehensive understanding of provider perspectives across different professional roles and telehealth experiences.

The survey was disseminated electronically to healthcare providers with a specialization in caring for YWH. Distribution channels included research networks such as the Adolescent Trials Network, Pediatric HIV/AIDS Cohort Studies Network, the International Maternal Pediatric Adolescent AIDS Clinical Trials Network, and the National AIDS Education and Training Center. Additionally, the survey was sent to mailing lists associated with the Association of Nurses in AIDS Care and the HIV Medicine Association and to the directors of the Centers for AIDS Research. The survey was also posted on selected social media sites and advertised at HIV-related conferences to broaden its reach. The survey collection period spanned from March 10, 2023, to June 20, 2023.

To compensate for their time and participation in the study, participants were offered a $25 gift card. This study was approved by The Committee for the Protection of Human Subjects (CPHS) at the University of Texas Health Science Center at Houston IRB (protocol HSC-SN-0659) and the Institutional Review Board for Baylor College of Medicine (protocol H-53567), which granted a waiver of written informed consent. Participants were informed about the purpose of the study, and consent was implied by their voluntary completion of the REDCap^29^ survey.

Upon completing data collection, all responses were compiled and reviewed to ensure accuracy and integrity. A total of 65 surveys were excluded as ineligible due to not meeting inclusion criteria. An additional 18 surveys were marked invalid due to incomplete or nonsensical responses, and 26 surveys were flagged as fraudulent, including submissions with fabricated institutional emails and repeated finder’s fee referrals. To address these concerns, three independent team members conducted a comprehensive review of all responses, using embedded quality-check questions to detect imposters, careless responders, and automated entries. This rigorous process ensured that only valid, high-quality responses were retained for analysis. After exclusions, 156 completed surveys met inclusion criteria and were included in the final analytic dataset.

## Data Analysis

Quantitative data were analyzed using descriptive statistics, mean and standard deviations for continuous variables and frequencies and percentages for categorical variables, to summarize the demographics, provider types, and responses to closed-ended questions. Associations between categorical variables were evaluated using Chi-squared tests. All statistical analyses were performed using R software (version 4.4.1) and RStudio (version 2024.09.0+375). ^30^

The qualitative component was analyzed using Braun and Clarke’s reflexive thematic analysis (RTA), a method that emphasizes researcher subjectivity and meaning-making in the development of themes. ^31,32^ This approach aligns with a constructivist epistemology, recognizing that themes are actively generated rather than objectively “emerging” from data. ^33^ Analysis was conducted using MAXQDA software^34^ to enhance the rigor and depth of the analysis. The team systematically identified, analyzed, and reported patterns (themes) within the data. To reduce individual bias, three team members trained in qualitative analysis independently reviewed and coded the data to gain an in-depth understanding of the content. Then the data was coded again using MAXQDA software. The team members compared and discussed their independently generated codes to enhance rigor and reduce bias, working collaboratively to refine and merge codes where necessary. Codes were translated into themes, constantly referring to the original data to ensure that these themes accurately represent the collected information. This iterative process, moving back and forth between the dataset, coded extracts, and the developing themes, a hallmark of Braun and Clarke’s approach, is facilitated by the software’s ability to reorganize and reclassify data easily. ^31–33^

The full team, including multidisciplinary healthcare providers of YWH, met and reviewed the final themes, ensuring they fit with both the coded extracts and the entire quantitative dataset to determine whether the themes captured the nuances of the data. Discrepancies were reviewed and discussed before finalizing and naming the themes. Detailed code sheets documenting the evolution of codes into themes, including definitions, inclusion and exclusion criteria for each theme, and illustrative quotes from the data to ensure transparency and replicability of the analysis, providing a clear audit trail of the researchers’ analytical decisions and interpretations.

## Results

### Sample Characteristics

A total of 156 HIV care providers completed the survey. The majority identified as female (77.6%, n=121) and non-Hispanic (87.2%, n=136), with 46.2% (n=72) aged between 40 and 59 years. Most respondents were prescribing providers (71.2%, n=112), including MDs, DOs, and advanced practice providers (APPs) like APRNs, NPs, and PAs; the remaining 28.8% (n=44) were nurses without prescriptive authority (e.g., RNs and MSNs). Providers primarily practiced in urban settings (83.3%, n=130), followed by suburban (13.5%, n=21) and rural areas (3.2%, n=5). Nearly half (44.9%, n=70) had more than 25 years of experience in HIV care. Importantly, over half of providers (51.9%, n=81) reported that 25% or more of their patients were youth aged 16–29 living with HIV, highlighting a strong youth care focus among the sample (Table 2).

**Table 2.**
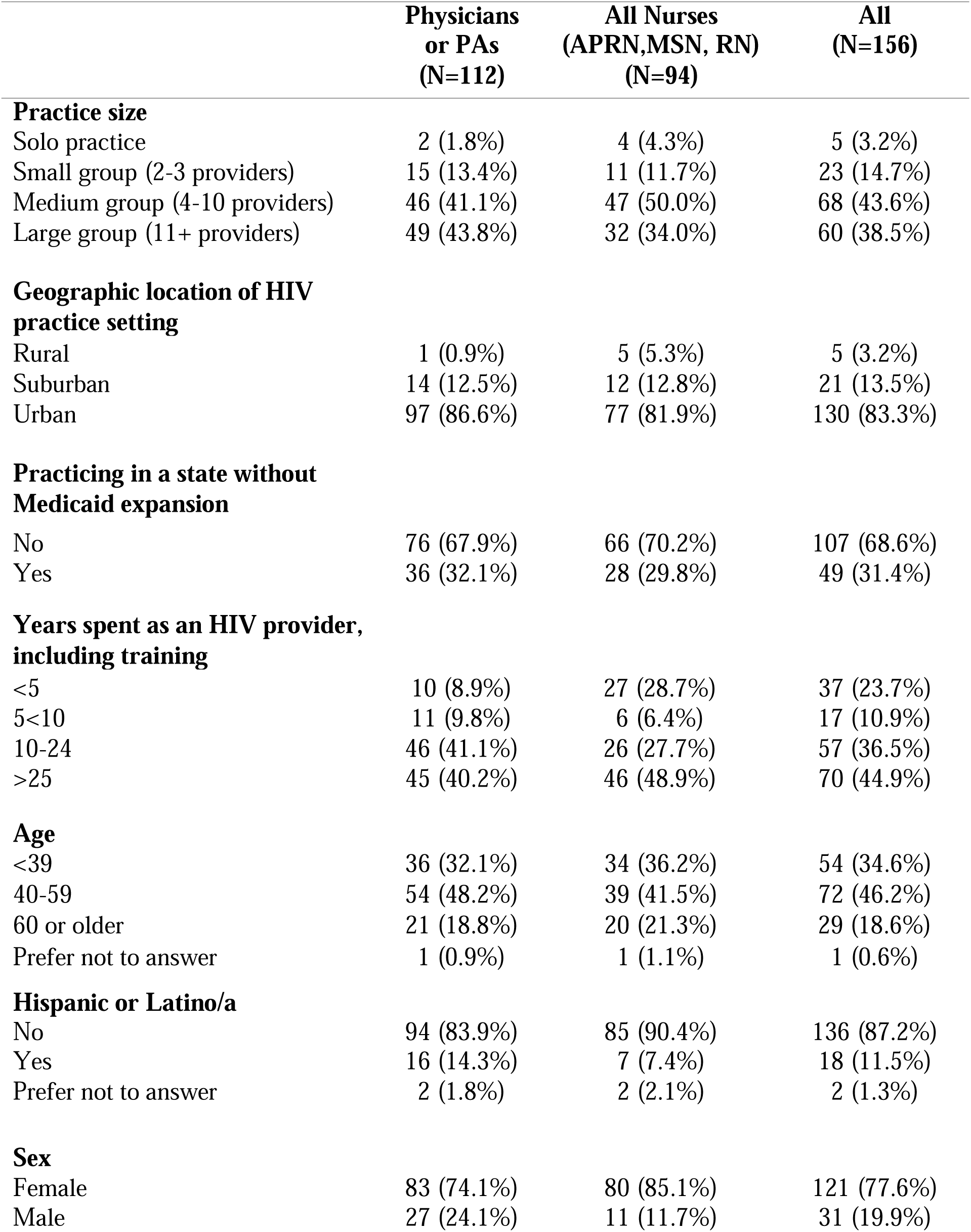
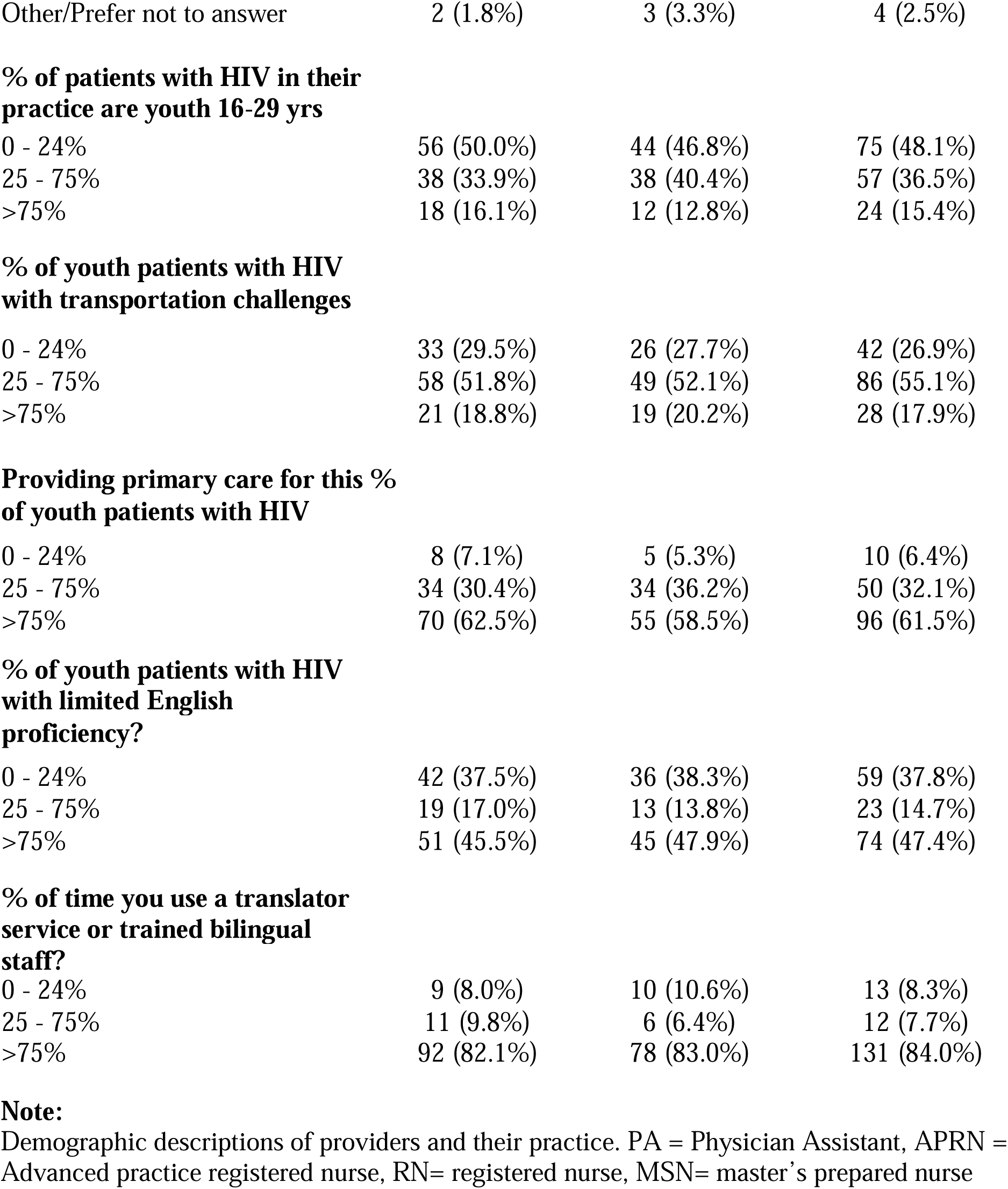
Demographics of HIV Care Providers and Practice Descriptions.

|  | **Physicians**<br>**or PAs**<br>**(N=112)** | **All Nurses**<br>**(APRN,MSN, RN)**<br>**(N=94)** | **All**<br>**(N=156)** |
| --- | --- | --- | --- |
| **Practice size** |  |  |  |
| Solo practice | 2 (1.8%) | 4 (4.3%) | 5 (3.2%) |
| Small group (2-3 providers) | 15 (13.4%) | 11 (11.7%) | 23 (14.7%) |
| Medium group (4-10 providers) | 46 (41.1%) | 47 (50.0%) | 68 (43.6%) |
| Large group (11+ providers) | 49 (43.8%) | 32 (34.0%) | 60 (38.5%) |
| **Geographic location of HIV**<br>**practice setting** |  |  |  |
| Rural | 1 (0.9%) | 5 (5.3%) | 5 (3.2%) |
| Suburban | 14 (12.5%) | 12 (12.8%) | 21 (13.5%) |
| Urban | 97 (86.6%) | 77 (81.9%) | 130 (83.3%) |
| **Practicing in a state without**<br>**Medicaid expansion** |  |  |  |
| No | 76 (67.9%) | 66 (70.2%) | 107 (68.6%) |
| Yes | 36 (32.1%) | 28 (29.8%) | 49 (31.4%) |
| **Years spent as an HIV provider,**<br>**including training** |  |  |  |
| <5 | 10 (8.9%) | 27 (28.7%) | 37 (23.7%) |
| 5<10 | 11 (9.8%) | 6 (6.4%) | 17 (10.9%) |
| 10-24 | 46 (41.1%) | 26 (27.7%) | 57 (36.5%) |
| >25 | 45 (40.2%) | 46 (48.9%) | 70 (44.9%) |
| **Age** |  |  |  |
| <39 | 36 (32.1%) | 34 (36.2%) | 54 (34.6%) |
| 40-59 | 54 (48.2%) | 39 (41.5%) | 72 (46.2%) |
| 60 or older | 21 (18.8%) | 20 (21.3%) | 29 (18.6%) |
| Prefer not to answer | 1 (0.9%) | 1 (1.1%) | 1 (0.6%) |
| **Hispanic or Latino/a** |  |  |  |
| No | 94 (83.9%) | 85 (90.4%) | 136 (87.2%) |
| Yes | 16 (14.3%) | 7 (7.4%) | 18 (11.5%) |
| Prefer not to answer | 2 (1.8%) | 2 (2.1%) | 2 (1.3%) |
| **Sex** |  |  |  |
| Female | 83 (74.1%) | 80 (85.1%) | 121 (77.6%) |
| Male | 27 (24.1%) | 11 (11.7%) | 31 (19.9%) |
| Other/Prefer not to answer | 2 (1.8%) | 3 (3.3%) | 4 (2.5%) |
| **% of patients with HIV in their practice are youth 16-29 yrs** |  |  |  |
| 0 - 24% | 56 (50.0%) | 44 (46.8%) | 75 (48.1%) |
| 25 - 75% | 38 (33.9%) | 38 (40.4%) | 57 (36.5%) |
| >75% | 18 (16.1%) | 12 (12.8%) | 24 (15.4%) |
| **% of youth patients with HIV with transportation challenges** |  |  |  |
| 0 - 24% | 33 (29.5%) | 26 (27.7%) | 42 (26.9%) |
| 25 - 75% | 58 (51.8%) | 49 (52.1%) | 86 (55.1%) |
| >75% | 21 (18.8%) | 19 (20.2%) | 28 (17.9%) |
| **Providing primary care for this % of youth patients with HIV** |  |  |  |
| 0 - 24% | 8 (7.1%) | 5 (5.3%) | 10 (6.4%) |
| 25 - 75% | 34 (30.4%) | 34 (36.2%) | 50 (32.1%) |
| >75% | 70 (62.5%) | 55 (58.5%) | 96 (61.5%) |
| **% of youth patients with HIV with limited English proficiency?** |  |  |  |
| 0 - 24% | 42 (37.5%) | 36 (38.3%) | 59 (37.8%) |
| 25 - 75% | 19 (17.0%) | 13 (13.8%) | 23 (14.7%) |
| >75% | 51 (45.5%) | 45 (47.9%) | 74 (47.4%) |
| **% of time you use a translator service or trained bilingual staff?** |  |  |  |
| 0 - 24% | 9 (8.0%) | 10 (10.6%) | 13 (8.3%) |
| 25 - 75% | 11 (9.8%) | 6 (6.4%) | 12 (7.7%) |
| >75% | 92 (82.1%) | 78 (83.0%) | 131 (84.0%) |

**Note:**
Demographic descriptions of providers and their practice. PA = Physician Assistant, APRN = Advanced practice registered nurse, RN= registered nurse, MSN= master's prepared nurse

Although no differences reached statistical significance, several trends emerged between nurses and physicians/PAs. Nurses were slightly more likely to report over 25 years of experience in HIV care (48.9% vs. 40.2%) and were more frequently based in rural settings (5.3% vs. 0.9%). While the majority of both groups identified as female, this was more pronounced among nurses (85.1%) compared to physicians/PAs (74.1%) (p = 0.078) (Table 2).

### Benefits and Barriers of Telehealth for YWH

Most providers in this study reported that telehealth may improve access to care (80.7%, n=117), that it will play a role in the future of HIV care (88.5%, n=138), and that telehealth improved daily workflow and efficiency in HIV care (Table 3). Additionally, a majority of the sample (80.7%, n=117), supported expanding telehealth services for YWH. Providers identified several other key benefits, including greater scheduling flexibility for patients (36.6%, n=53) and improved convenience for providers (40.0%, n=58). When asked to rank patient-specific benefits, the most frequently selected top benefit was decreased practical barriers to care for patients, such as transportation or childcare (66.9%, n=97). Providers reported an ideal practice of median 25% visits occur via Telehealth (with mean percentage 32.1% of visits. with a preference for video-based modalities rather than audio-only or text-based options, as demonstrated by most providers agreeing or strongly agreeing with the statement that video visits should be expanded for YWH (n=117, 80.7%).

**Table 3.** Ranked Telehealth Benefits and Barriers by Providers.

| Category | Top 3 Ranked | Bottom 3 Ranked |
| --- | --- | --- |
| Patient benefits | 1. Decreases barriers to care (i.e. transportation, childcare, etc.) n=97 (66.9%)<br>2. Patients prefer flexibility n=53 (36.6%)<br>3. Saved time for patients n=44 (33.3%) | 4. Protects privacy n=38 (26.2%)<br>5. Protects the health of patients n=47 (32.4%)<br>6. Increased ability to build patient-provider trust n=53 (36.6%) |
| Provider benefits | 1. Convenient for providers n=58 (40.0%)<br>2. Improves clinic efficiency n=53 (36.6%)<br>3. Fewer no-show/canceled appointments n=40 (27.6%) | 4. Protects patients/staff n=33 (22.8%)<br>5. Increases patient-provider trust n=63 (43.4%)<br>6. Favorable compensation n=64 (44.1%) |
| Patient barriers | 1. Technical difficulties for the patient n=51 (35.2%)<br>2. Inability for patients to complete paperwork/meet with support staff n=31 (21.4%)<br>3. Access to technology n=23 (15.9%) | 4. Concerns about patient privacy n=31 (21.4%)<br>5. Decreased ability to build trust with provider n=39 (26.9%)<br>6. Unfavorable insurance coverage n=54 (37.2%) |
| Provider barriers | 1. Challenges obtaining labs, STI tests, or other clinical tests n=55 (37.9%)<br>2. Inability to perform a complete physical exam n=38 (26.2%)<br>3. Barrier to providing vaccines n=40 (27.6%) | 4. Obtaining vital signs n=33 (22.8%)<br>5. Patient may be distracted or not in an appropriate setting n=36 (24.8%)<br>6. Challenges in building patient-provider trust n=59 (40.7%) |

**Note:** Describes the benefits of in-person vs. telehealth, as reported by providers (n=145).

While 45.5% of providers (n=71) were neutral about the impact of telehealth on patient engagement, 39.7% (n=62) believed it enhanced engagement and adherence, and 14.7% (n=23) believed it had a negative impact. Providers were more likely to view telehealth as appropriate for youth already engaged in care (37.8%, n=59) compared to those less engaged (7.1%, n=11). Nonetheless, most reported offering telehealth regardless of a patient’s engagement level (Table 4).

**Table 4.**
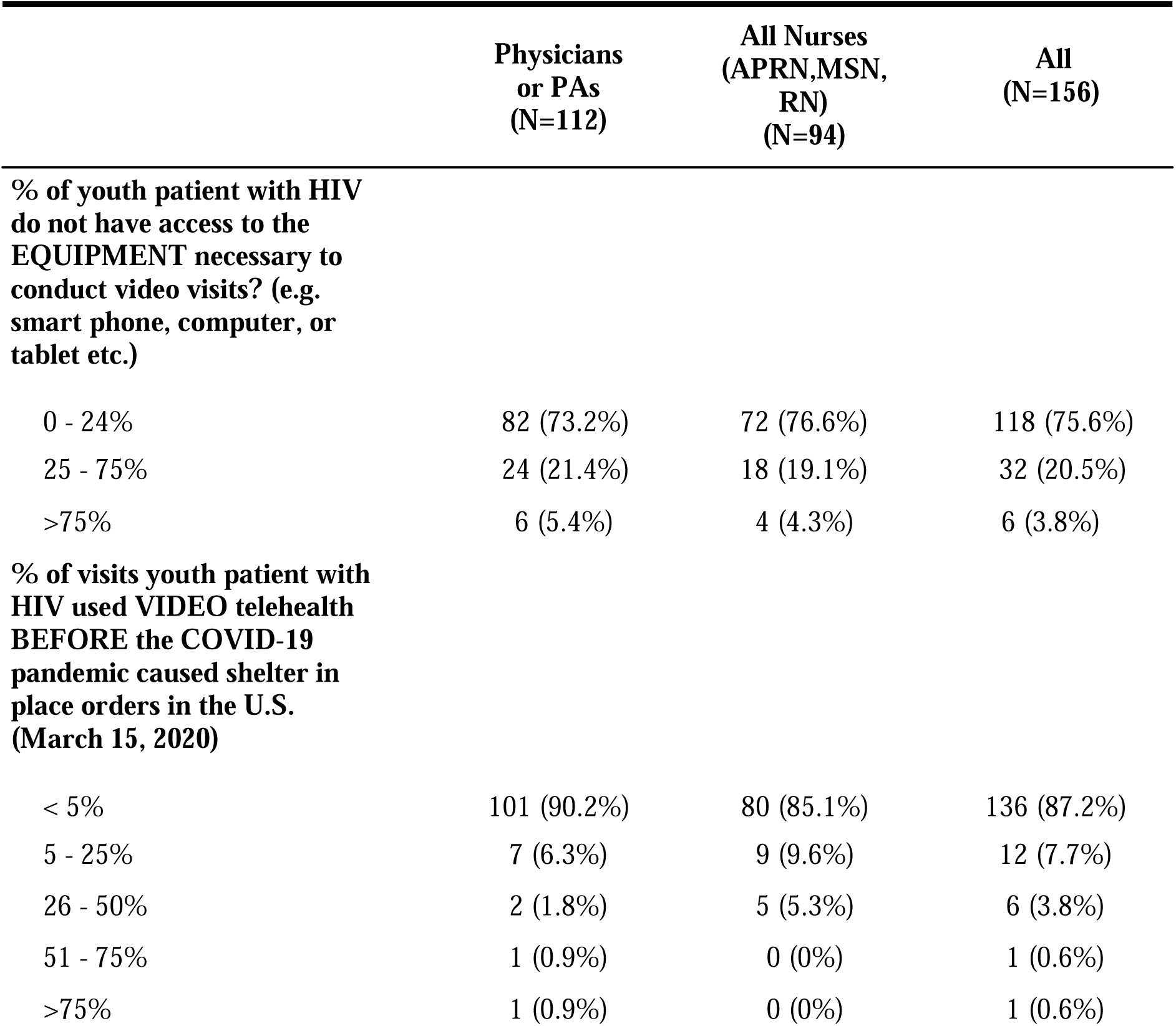
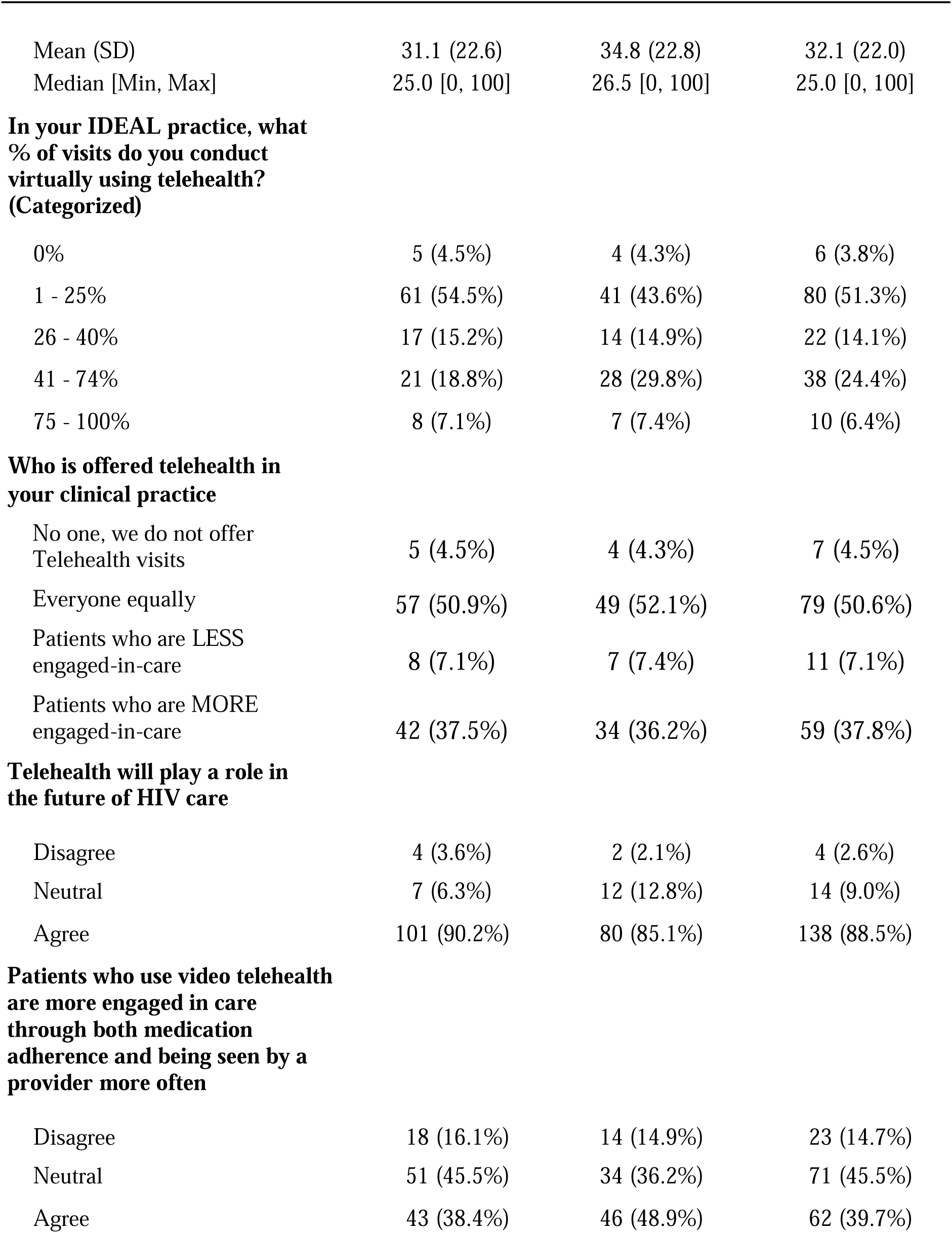
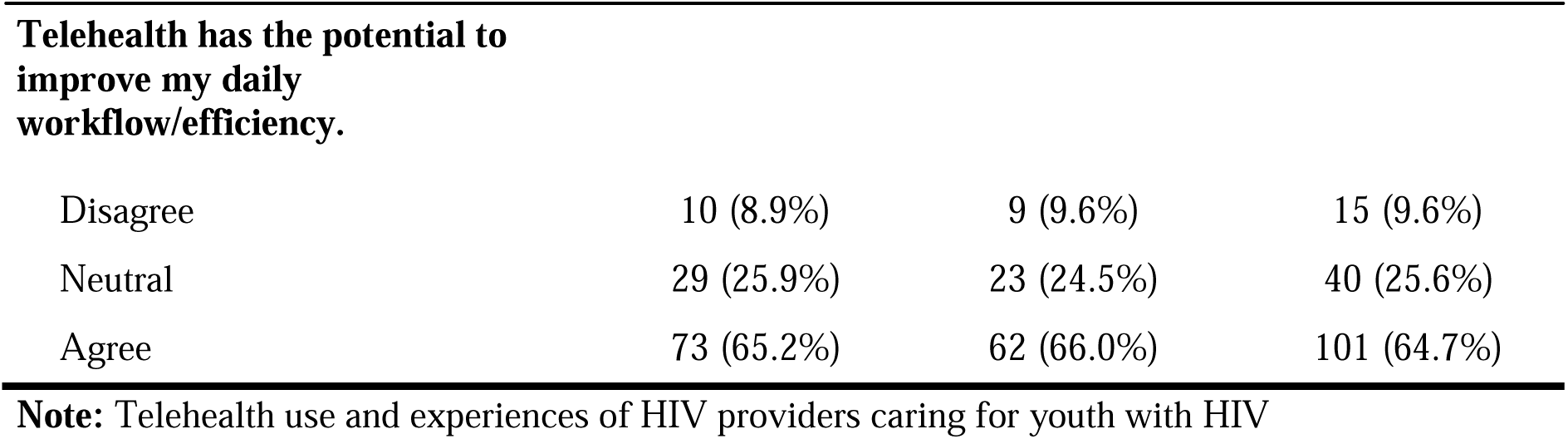
Telehealth Use and Preferences by Provider Type.

#### Technology

Despite its advantages, providers identified significant barriers to telehealth implementation for both patients and providers (Table 3). The most frequently cited challenge was patient-side technical difficulties, such as unreliable internet, lack of access to a smartphone or laptop, or limited digital literacy, with 35.2% (n=51) of providers ranking this as the top patient barrier. Additionally, 26.9% (n=30) reported that more than 25% of their youth patients lacked the necessary equipment for video visits. Only 26.2% (n=38) of providers felt that video visits allowed patients to feel more open, and just 17.9% (n=26) agreed that video visits improved the overall quality of care (Table 5).

**Table 5.**
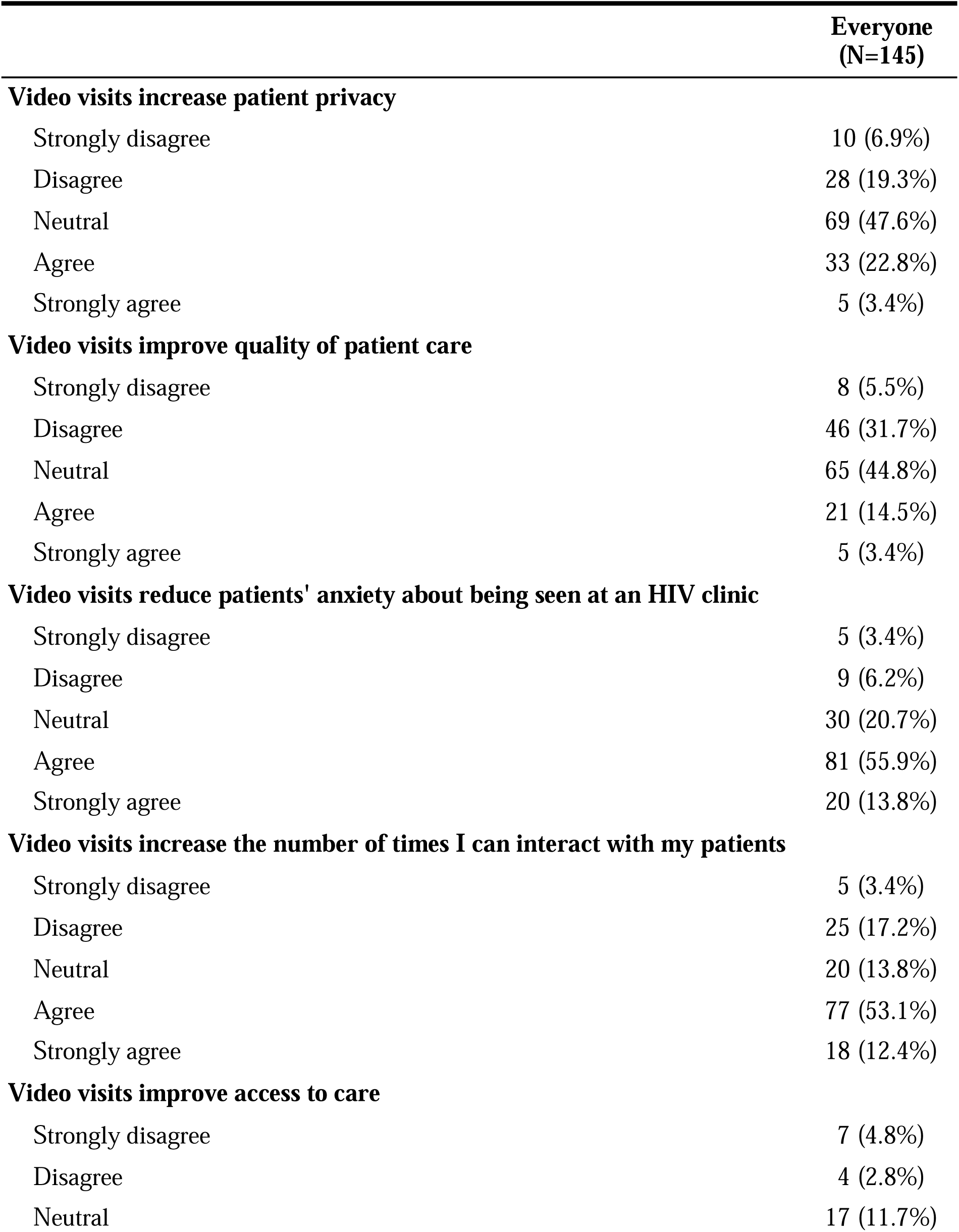
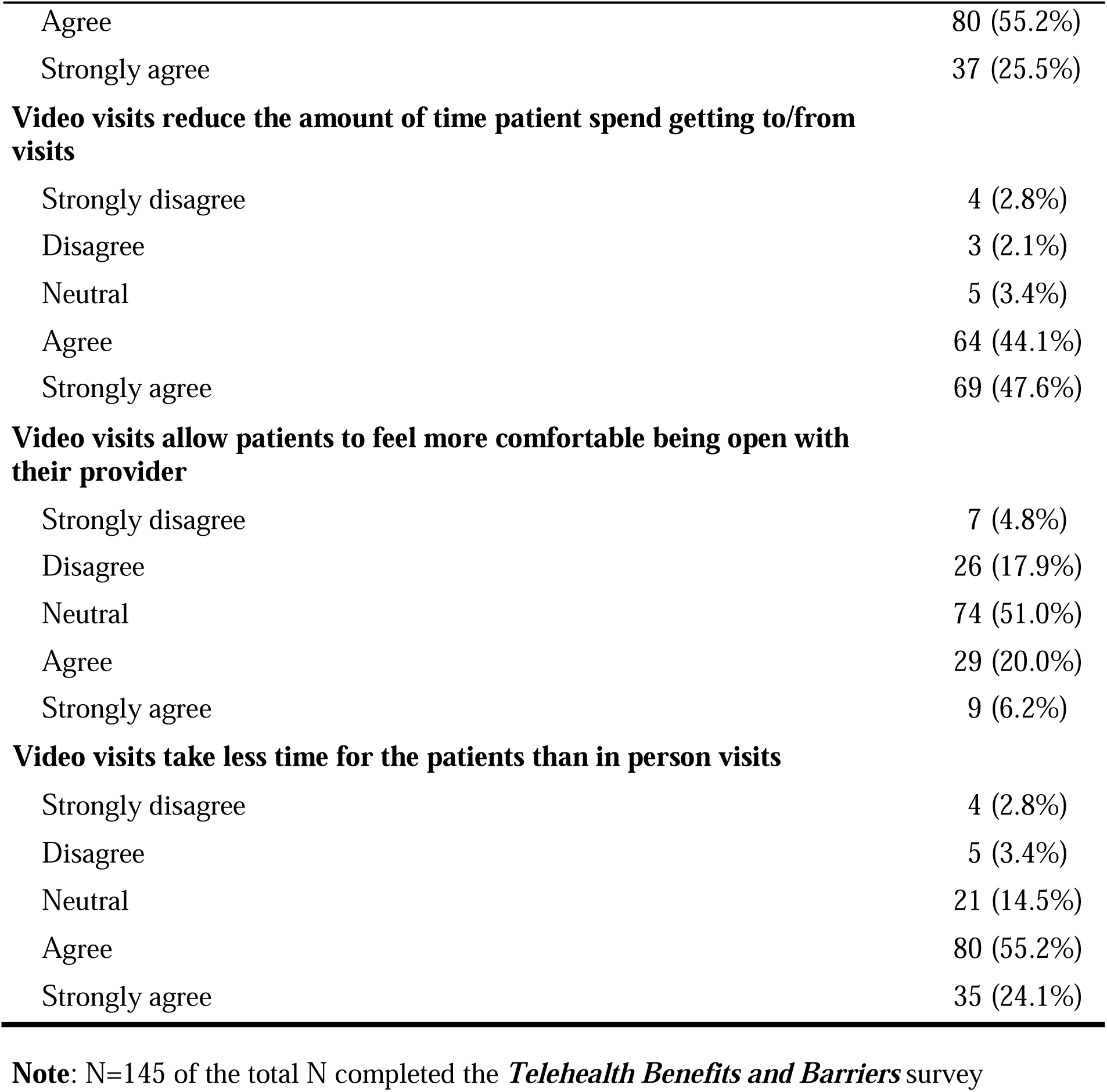
Perceptions of Video Telehealth: Benefits and Barriers.

|  | **Everyone**<br>**(N=145)** |
| --- | --- |
| **Video visits increase patient privacy** |  |
| Strongly disagree | 10 (6.9%) |
| Disagree | 28 (19.3%) |
| Neutral | 69 (47.6%) |
| Agree | 33 (22.8%) |
| Strongly agree | 5 (3.4%) |
| **Video visits improve quality of patient care** |  |
| Strongly disagree | 8 (5.5%) |
| Disagree | 46 (31.7%) |
| Neutral | 65 (44.8%) |
| Agree | 21 (14.5%) |
| Strongly agree | 5 (3.4%) |
| **Video visits reduce patients' anxiety about being seen at an HIV clinic** |  |
| Strongly disagree | 5 (3.4%) |
| Disagree | 9 (6.2%) |
| Neutral | 30 (20.7%) |
| Agree | 81 (55.9%) |
| Strongly agree | 20 (13.8%) |
| **Video visits increase the number of times I can interact with my patients** |  |
| Strongly disagree | 5 (3.4%) |
| Disagree | 25 (17.2%) |
| Neutral | 20 (13.8%) |
| Agree | 77 (53.1%) |
| Strongly agree | 18 (12.4%) |
| **Video visits improve access to care** |  |
| Strongly disagree | 7 (4.8%) |
| Disagree | 4 (2.8%) |
| Neutral | 17 (11.7%) |
| Agree | 80 (55.2%) |
| Strongly agree | 37 (25.5%) |
| **Video visits reduce the amount of time patient spend getting to/from visits** |  |
| Strongly disagree | 4 (2.8%) |
| Disagree | 3 (2.1%) |
| Neutral | 5 (3.4%) |
| Agree | 64 (44.1%) |
| Strongly agree | 69 (47.6%) |
| **Video visits allow patients to feel more comfortable being open with their provider** |  |
| Strongly disagree | 7 (4.8%) |
| Disagree | 26 (17.9%) |
| Neutral | 74 (51.0%) |
| Agree | 29 (20.0%) |
| Strongly agree | 9 (6.2%) |
| **Video visits take less time for the patients than in person visits** |  |
| Strongly disagree | 4 (2.8%) |
| Disagree | 5 (3.4%) |
| Neutral | 21 (14.5%) |
| Agree | 80 (55.2%) |
| Strongly agree | 35 (24.1%) |

**Note:** N=145 of the total N completed the *Telehealth Benefits and Barriers* survey

#### Access to Care & Engagement in Care

Although most providers believed that telehealth improved access and convenience, 66.2% (n=103) felt that communication was more effective in person. Providers acknowledged that telehealth allowed for more frequent check-ins and timely adherence support, but only 39.7% (n=62) agreed that it meaningfully enhanced engagement, while 45.5% (n=71) were neutral and 14.7% (n=23) disagreed.

Table 4 compares responses between providers with a primarily medical model background (physicians and PAs) and those with a nursing background (nurses and APRNs). Perceptions of telehealth’s impact on engagement and care outcomes were mixed across both groups. Overall, 45.5% of providers were neutral about its influence on youth engagement, while 39.7% believed youth who used telehealth were more engaged and adherent, and 14.7% believed they were less engaged. Nurses were slightly more likely than physicians/PAs to view telehealth as enhancing engagement (48.9% vs. 38.4%), while physicians/PAs were more likely to express neutrality (45.5% vs. 36.2%). Although these differences were not statistically significant, they suggest subtle distinctions in perception by provider type. Regardless of background, the overwhelming majority agreed that telehealth would continue to play a role in the future of HIV care delivery, indicating widespread support for its continued integration.

#### Communication and Care Content

Providers reported that core components of care, such as medication adherence, lab results, personal life stressors, substance use, mental health, and sexual health, were addressed in both telehealth and in-person encounters (Table 3). However, there were clear differences in how frequently sensitive topics were discussed. Providers were significantly less likely to report discussing psychosocial and sensitive topics “often” during telehealth visits. For example, home and work life were discussed often in 78.8% of telehealth visits versus 94.9% of in-person visits (p < 0.0001). Similar differences were noted for substance use (72.1% vs. 92.3%, p < 0.0001), mental health (81.4% vs. 92.9%, p = 0.0008), and sexual health (77.9% vs. 93.6%, p < 0.0001). These differences remained statistically significant after applying a Bonferroni correction (adjusted p < 0.0083), underscoring a consistent pattern of reduced depth in virtual visits.

#### Clinical Limitations and Care Quality

Providers reported that telehealth was less effective for complex clinical encounters. Although telehealth was considered useful for routine follow-up care, including medication management and adherence counseling, many providers highlighted the limitations in conducting physical examinations or collecting laboratory specimens remotely. Even when providers covered essential topics such as medication adherence and test results, they were significantly less likely to report discussing psychosocial or sensitive topics during telehealth visits compared to in-person visits (Table 6). Providers were less likely to discuss home and work life (78.8% telehealth vs. 94.9% in-person, p < 0.0001), substance use (72.1% vs. 92.3%, p < 0.0001), mental health (81.4% vs. 92.9%, p = 0.0008), and sexual health (77.9% vs. 93.6%, p < 0.0001) during remote encounters (Table 6). These differences remained statistically significant after applying a Bonferroni correction. ^35^

**Table 6.** Frequency of Health Topics Discussed During In-Person vs. Telehealth Visits.

| *Question: How often do you talk about the following in an in-person visit and in a telehealth visit?* |  |  |  |  |
| --- | --- | --- | --- | --- |
| **Question:** | **Answers<sup>1</sup>** | **Visit Type** |  | **P-value** |
|  |  | In-person visit<br>(N=156) | Telehealth<sup>2</sup><br>(N=312) |  |
| Adherence to medications | <=Sometimes | 3 (1.9%) | 19 (6.1%) | 0.0614 |
|  | >=Often | 153 (98.1%) | 293 (93.9%) |  |
| Test results<br>(e.g. labs, imaging) | <=Sometimes | 4 (2.6%) | 22 (7.1%) | 0.05355 |
|  | >=Often | 152 (97.4%) | 290 (92.9%) |  |
| Home and work life (e.g. housing status, life stressors) | <=Sometimes | 8 (5.1%) | 66 (20.2%) | <0.0001* |
|  | >=Often | 148 (94.9%) | 246 (78.8%) |  |
| Substance use (e.g. tobacco, alcohol, drug use) | <=Sometimes | 12 (7.7%) | 87 (27.9%) | <0.0001* |
|  | >=Often | 144 (92.3%) | 225 (72.1%) |  |
| Mental health | <=Sometimes | 11 (7.1%) | 58 (18.6%) | 0.0008* |
|  | >=Often | 145 (92.9%) | 254 (81.4%) |  |
| Sexual health (e.g. STIs, PrEP, birth control, condom use) | <=Sometimes | 10 (6.4%) | 69 (21.1%) | <0.0001* |
|  | >=Often | 146 (93.6%) | 243 (77.9%) |  |

Notes:
<sup>1</sup>Original response options were: “Never,” “Rarely,” “Sometimes,” “Often,” and “Always.” Responses were collapsed into two categories for analysis: ≤ Sometimes includes “Never,” “Rarely,” or “Sometimes,” and ≥ Often includes “Often” or “Always.”
<sup>2</sup>Telehealth includes audio and video telehealth combined
\*Significant after the Bonferroni multiple testing adjustment ( $p=0.05/6 = 0.0083$ )

Providers shared mixed concerns about the effects of telehealth on relational aspects of health care, such as quality of interaction, the experience of social stigma, or trust within the patient-provider relationship. While 69.7% (n=101) of providers agreed that video visits helped reduce patient anxiety about being seen at an HIV clinic, only 65.5% (n=95) felt telehealth improved opportunities for patient interaction (Table 5).

#### Policy & Reimbursement

Another critical barrier involved uncertainty regarding long-term reimbursement for telehealth. While federal and state-level regulations during the COVID-19 pandemic expanded reimbursement, many providers expressed concern about the sustainability of these policies. More than half of respondents ranked inadequate compensation or lack of insurance coverage among the top three provider-side barriers (Table 3). These concerns were especially pronounced among providers working in smaller, resource-limited clinics.

### Qualitative Findings

Providers’ free-text responses largely echoed the quantitative survey findings and offered critical nuance on their experiences using telehealth with YWH. Reflexive thematic analysis revealed three central themes: ***Technology***, ***Access & Engagement***, and ***Intimacy***. These themes help contextualize and deepen interpretation of the survey data, supporting best practices for integration in mixed-methods research.

#### Technology

Consistent with survey results showing technical challenges as a leading barrier (Table 3), providers described unstable internet, limited access to devices, and low digital literacy among patients as frequent obstacles to effective care. As one family nurse practitioner with 5– 25 years of experience explained, “Telehealth helps keep patients with transportation, location, and work barriers engaged in care.” However, others noted that infrastructure gaps undermined access, particularly in rural areas. “I see these limitations primarily on the patient-side, with poor cellular connectivity in rural portions of our state,” shared a physician with 5–25 years of experience. These qualitative perspectives reinforce the survey findings that one in four providers reported more than 25% of their youth patients lacked the necessary equipment for video visits.

#### Access & Engagement

Many providers emphasized that telehealth could support engagement by reducing barriers and increasing patient autonomy, echoing quantitative data where 39.7% of respondents believed telehealth users were more engaged and adherent (Table 4). One nurse practitioner with under 5 years of experience shared, “Electronic health care allows patients to feel more empowered in their health outcomes.” Similarly, a physician reported that telehealth “has been excellent in getting [patients] back to clinic and engaging in their own health.” Several providers noted that electronic tools enhanced patient engagement, particularly through real-time access to appointments and lab results. As one nurse practitioner with under 5 years of experience shared, “I think patients really appreciate the ability to see their appointments and results in real time.” Another provider noted that the *“*patient portal is useful to some patients as a way to communicate concerns/questions that they might forget during their visit” (physician, 5–25 years of experience). These tools were viewed as supporting autonomy and helping youth stay engaged between visits.

However, this optimism was tempered by concerns that some youth may disengage without strong self-management skills. “Although we do everything we can to make it easy for patients to go in for labwork…it takes a lot of reminders and sometimes doesn’t happen,” shared a physician. Another provider noted the distractions in non-private settings: “I have asked the patient if they were comfortable talking where they were… only to find out they were in the supermarket or laundromat and definitely not paying attention. I worried about the effectiveness of the visit and about HIPAA” (advanced practice nurse, over 25 years of experience). These reflections mirror quantitative data showing that providers were significantly less likely to discuss sensitive topics like mental health or sexual health via telehealth (Table 6).

#### Intimacy

Trust, comfort, and emotional connection, key to sensitive HIV care, emerged as central concerns in telehealth delivery. While quantitative data showed only 26.2% of providers believed video visits helped patients be more open (Table 5), qualitative responses offered a more nuanced view. Several providers described feeling limited in their ability to build intimacy virtually, particularly during emotionally or clinically complex conversations. As one provider described, “It is a little more difficult to talk about more sensitive topics virtually, especially if there are connection issues” (physician, 5–25 years of experience).

Yet others found that the privacy of home allowed for more honest disclosure. A physician with over 25 years of experience remarked, “I’ve had several patients disclose information during a call that they had not previously disclosed in person.” This contrast mirrors the quantitative ambivalence on communication quality, where 66.2% felt in-person visits were better, but many acknowledged telehealth’s convenience and its role in reducing stigma (Table 5).

Providers also emphasized the need for enhanced training and support. A physician assistant with under 5 years of experience called for “additional training on best methods for conducting virtual visits, especially when assessing mental health concerns.” Another physician suggested patient-side training: “how to find and turn on the microphone and camera, etc.” These calls align with broader findings in the survey that providers see potential in telehealth but need structured support to optimize its use.

## Discussion

Findings from this mixed-methods study suggest that providers generally view telehealth as a valuable adjunct to traditional care. In both survey and qualitative responses, providers noted that telehealth reduced logistical barriers, such as transportation and scheduling conflicts, and facilitated more frequent patient check-ins. This aligns with broader research documenting telehealth’s utility for improving access among hard-to-reach populations. However, the benefits of telehealth were tempered by concerns about access to technology and variability in patient readiness. Over one-third of providers cited patient-side technical issues as a major barrier, and nearly 27% reported that a quarter or more of their YWH patients lacked the necessary equipment for video visits. These patterns may also vary by provider background; nurses in this sample were more likely than physicians to work in rural settings and to report over 25 years of experience, which could shape how they approach telehealth implementation and patient engagement. These findings point to the need for digital equity initiatives, including device access and broadband infrastructure, to ensure inclusive care delivery.

Providers noted that telehealth helped mitigate logistical barriers for YWH, including transportation and scheduling challenges. Digital tools such as patient access to electronic medical records (EMRs) and patient portals were identified as valuable for promoting engagement, enabling youth to access health information, manage appointments, and communicate with providers. In qualitative responses, providers emphasized that these platforms can empower YWH to take greater ownership of their health. However, they also cautioned that some youth may struggle to stay engaged without strong self-management skills. Overall, providers viewed telehealth as a promising tool to enhance access, autonomy, and engagement in care, while acknowledging that fostering intimacy and trust in virtual settings may require intentional strategies. ^36^

Trust and communication emerged as central concerns when delivering care virtually. Although many providers felt telehealth allowed for greater privacy and reduced stigma, they were significantly less likely to discuss sensitive topics such as mental health, sexual health, or substance use during telehealth visits compared to in-person care. These challenges underscore the importance of enhancing provider training in virtual communication techniques and building comfort in addressing sensitive issues remotely.

Interestingly, some providers described unexpected moments of enhanced intimacy during virtual visits, suggesting that for certain patients, the familiarity of home settings may foster openness. This complexity highlights that communication quality in telehealth is not universally better or worse, it is context-dependent and influenced by both provider skill and patient environment. Such concerns reflect those documented in previous literature, which highlight the limitations of telehealth in performing comprehensive physical assessments and fostering the depth of rapport necessary for discussing sensitive topics, such as mental health or sexual behavior. ^6,37^

Providers also emphasized that telehealth alone is insufficient for high-quality HIV care. Many noted its limitations for physical assessments, laboratory testing, and addressing psychosocial needs. Across both quantitative and qualitative data, a hybrid care model, combining telehealth with periodic in-person visits, emerged as the preferred approach. This model was seen as optimizing access while maintaining the depth of care required for complex patients. Providers who integrated telehealth with periodic in-person visits reported more positive outcomes, indicating that a hybrid model may be more effective in managing the complex care needs of YWH.

Finally, providers identified a strong need for structural and training support to optimize telehealth. Suggestions included developing best practices for virtual trust-building, ensuring patient privacy, and standardizing workflows for managing laboratory tests and mental health evaluations. Investing in provider and patient training could strengthen the quality of virtual care and reduce disparities in telehealth engagement.

## Limitations

This study has several limitations. First, the sample was limited to providers in the United States who responded to an online survey, which may introduce selection bias; those more comfortable with technology or with stronger opinions about telehealth may have been more likely to participate. While diverse in discipline, geography, and years of experience, the sample may not represent all provider perspectives, particularly from rural or under-resourced settings where telehealth infrastructure is more limited. Second, the data were self-reported and may be subject to recall or social desirability bias, especially regarding sensitive topics such as provider comfort or patient disclosure during virtual visits. Additionally, although the mixed-methods design allowed for nuanced exploration of telehealth experiences, the qualitative data were drawn from open-ended survey responses rather than in-depth interviews or focus groups, which may limit the depth of insight. Finally, the study was conducted in the post-acute phase of the COVID-19 pandemic, during which telehealth policies and workflows were in flux. Provider experiences and attitudes may shift over time as regulatory, reimbursement, and technological landscapes continue to evolve. Future longitudinal and implementation-focused studies are needed to assess how telehealth practices and impacts change in more stable healthcare environments.

## Conclusion

This study highlights both the promise and the complexity of telehealth in HIV care for youth. Providers widely endorsed telehealth as a tool to enhance access, reduce logistical barriers, and support continuity of care, especially for youth who face challenges with transportation, school, or work. However, this optimism was tempered by persistent barriers, including unequal access to technology, limitations in addressing sensitive topics, and concerns about diminished trust and intimacy in virtual settings. Addressing these barriers will be critical to ensure that telehealth can meet the comprehensive needs of YWH, particularly in managing the multifaceted nature of their care. To realize the full potential of telehealth for youth with HIV, targeted training in virtual communication, investment in digital infrastructure, and clear clinical protocols for hybrid care are essential. As healthcare systems evolve, centering the voices and needs of providers and young people will be key to designing effective, responsive, and youth-affirming care models.

## Data Availability

All data produced in the present study are available upon reasonable request to the authors.

## Acknowledgements

Litty Koshy, Markeda Wade, Association of Nurses in AIDS Care (ANAC), AIDS Education and Training Centers (AETC), the Pediatric HIV/AIDS Cohort Study (PHACS), the International Maternal Pediatric Adolescent AIDS Clinical Trials Network (IMPAACT), and Adolescent Medicine Trials for HIV (ATN) networks for disseminating the survey.

